# Sex-stratified genomic structural equation models of posttraumatic stress inform PTSD etiology

**DOI:** 10.1101/2023.09.01.23294954

**Authors:** Ashley Moo-Choy, Murray B Stein, Joel Gelernter, Frank R Wendt

## Abstract

Posttraumatic stress disorder (PTSD) affects 3.9%-5.6% of the worldwide population, with well-documented sex-related differences. While psychosocial and hormonal factors affecting sex differences in PTSD and posttraumatic stress (PTS) symptom etiology have been explored, there has been limited focus on genetic bases of these differences. Many symptom combinations may confer a PTSD diagnosis. We hypothesized that these symptom combinations have sex-specific patterns, the examination of which could inform etiological differences in PTSD genetics between males and females. To investigate this, we performed a sex-stratified multivariate genome-wide association study (GWAS) in unrelated UK Biobank (UKB) individuals of European ancestry. Using GWAS summary association data, genomic structural equation modeling was performed to generate sex-specific factor models using six indicator variables: trouble concentrating, feeling distant from others, irritability, disturbing thoughts, upset feelings, and avoidance of places/activities which remind the individual of a traumatic event. Models of male and female PTSD symptoms differed substantially (local standardized root mean square difference=3.12) and significantly (χ^2^(5)=28.03, p=3.6×10^-5^). Independent two-factor models best fit the data in both males and females; these factors were subjected to GWAS in each sex, revealing three genome-wide significant loci in females, mapping to *SCAND3, WDPCP*, and *FAM120A*. No genome-wide significant loci were identified in males. All four PTS factors (2 in males and 2 in females) were heritable (p<0.05): male PTS-f_1_ *h*^*2*^-SNP=1.85%, male PTS-f_2_ *h*^*2*^-SNP=1.47%, female PTS-f_1_ *h*^*2*^-SNP=3.87%, female PTS-f_2_ *h*^*2*^-SNP=3.53%. Male PTS-f_1_ was enriched for medication-related putative causal relationships (3.73-fold, p=0.032) while male PTS-f_2_ was enriched for body structure (2.84-fold, p=5.512×10^-7^) and cognitive (3.22-fold, p=0.009) putative causal relationships. In females, PTS-f_1_ was enriched for metabolic putative causal relationships (28.08-fold, p=0.035). By assessing the relationship between sex and PTSD symptoms, this study informs correlative and putatively causal etiological differences between males and females which support further investigation of sex differences in PTSD genetics.

## INTRODUCTION

Posttraumatic stress disorder (PTSD) affects an estimated 3.9% to 5.6% of the worldwide population (1). In individuals with severe mental illness, an estimated 30% also suffer from PTSD as a result of traumatic experiences (2). The Diagnostic and Statistical Manual of Mental Disorders, 5^th^ Edition (DSM-5) defines traumatic exposure as either experiencing, witnessing, and/or learning of a traumatic event involving friends or family, or by repetitive personal exposure, such as by first responders or police officers (3). Diagnosis of PTSD after trauma exposure requires an individual to experience intrusive (aka reexperiencing) memories of the traumatic event(s), hyperarousal, avoidance of reminders of the event(s), and negative cognitions and mood related to the trauma for a period of at least one month, resulting in significant distress and/or functional impairment.

Trauma exposure and PTSD are both heritable, meaning genetic factors explain a portion of trait variation in the population (4). Trauma exposure affects people differently based on sex, gender identity, and age of exposure (5,6). Women have at least two times higher lifetime risk of PTSD, in part due to exposure to different types of trauma than men (e.g., rape and intimate partner violence) and age at trauma exposure (e.g., childhood sexual abuse). There have been both biological (e.g. hormonal) and psychosocial hypotheses as to the cause of sex-based differences in PTSD onset, symptomatology, and treatment, but sex-based empirical biological data are lacking (4). Women are more likely to experience sexual and familial trauma at younger ages, while men are more likely to be exposed to trauma in adulthood. Conversely, some studies suggest that the increased prevalence of PTSD in women is due to the differing symptomatology of PTSD relative to men (7). For example, women report more re-experiencing symptoms than men, motivating the exploration of different etiologies and patterns of symptom clustering.

Previous genome-wide association studies (GWAS) of PTSD have appreciated these known sex differences in exposure to trauma and PTSD symptom presentation. The SNP-based heritability of PTSD in women is consistently non-zero and detectable even in early studies of PTSD with limited sample size (8,9). Conversely, studies in men required larger sample sizes to detect a significant SNP-based heritability estimate (8,9). Independent of sex and/or gender differences in PTSD presentation, PTSD genetics correlates with genetic factors influencing depression (10), anxiety (11), attention deficit hyperactivity disorder (12), schizophrenia (8), bipolar disorder (13), and other mental disorders. Prior reports postulate that various combinations of PTSD risk factors, including symptom patterns and severity, explain a large portion of sex differences in PTSD etiology (14). As we recognize the phenotypic differences in PTSD between males and females, we investigated whether we could identify a genetic basis of these differences.

DSM-5 criteria enable over 636,000 possible diagnostic combinations of PTSD symptoms, and prior work demonstrates substantial diagnostic overlap between patients (15,16). We hypothesized that PTSD symptoms overlap in a sex-specific manner and that this pattern of overlap informs sex-specific disorder etiology. We used genomic structural equations modelling (gSEM) to model multivariate genetic architecture of PTSD symptoms separately in males and females. We report sex-specific factor structures whose genetic architecture differentially overlaps with features of the human phenome that inform differences in PTSD etiology between males and females.

## METHODS

### Data Description and Factor Modeling

The UK Biobank (UKB) is a population-based cohort of >500,000 participants with deep phenotyping of lifestyle factors, mental and physical health outcomes, anthropometric measurements, and other traits. This study used GWAS summary data for posttraumatic stress (PTS) phenotypes from unrelated European ancestry participants adjusted for the first 20 genetic principal components, age, and age^2^. Sex was not included as a covariate due to the sex stratified design of this study.

gSEM models the multivariate genetic architecture of sets of traits considering their genetic covariance structure (17). Using gSEM, a multivariable GWAS was performed with six PTS traits based on their unique factor structure in males and females. The six factor indicators were questions from the UKB thoughts and feelings section of the online Mental Health Questionnaire and have been previously described by Davis et al. (Table S1) (18). Five questions asked about how often participants were bothered by: (i) *avoiding activities or situations because they remind them of stressors* (“Avoid”, N=117,868, 56% female), (ii) *feeling distant or cut off from others* (“Distant”; N=52,822; 63% female), (iii) *feeling irritable or having angry outbursts* (“Irritable”, N=52,816, 63% female), (iv) *repeated, disturbing memories, thoughts, or images of a stressful experience* (“Thoughts”, N=117,900, 56% female), and (v) *feeling very upset when something reminded them of a stressor* (“Upset”, N=117,893, 56% female). Participants responded to these questions with a rating from 0 =”not at all” to 4 =”extremely.” The sixth question asked about how bothered participants were by *trouble concentrating on things, such as reading or watching television*, over the last two weeks (“Concentrate”, N=117,899, 56% female). Participants responded to this question with a rating from 1 =”not at all” to 4 =”nearly every day.”

Multivariable linkage disequilibrium score regression (LDSC) was used to obtain genetic covariance and sampling matrices using a European ancestry linkage disequilibrium reference panel from the 1000 Genomes Project. Factor modeling used diagonally weighted least squares estimation and promax rotation. Exploratory factor analysis (EFA) was used to evaluate a common factor and a two-factor structure for the six traits in males and females. From EFA, confirmatory factor analysis (CFA) was performed using all indicator traits with factor loadings > 0.3. Four model fit statistics were compared to assess the suitability of a one-factor versus two-factor model: chi-squared (χ^_2_^), comparative fit index (CFI), Akaike information criterion (AIC), and standardized root mean square residual (SRMR). The χ^2^ statistic indexes whether modelled genetic covariance differs from an empirical matrix. CFI tests whether the proposed model fits better than a model that assumes all indicators are heritable but uncorrelated. AIC measures relative model fit and may be used to compare multiple models. SRMR measures approximate model fit calculated as the standardized root mean square difference between implied and observed correlations among covariance matrices. Each fit statistic has strengths and weaknesses, so we considered several features in deciding the best model. When comparing one- and two-factor models, the superior model in our study had a low AIC value, low SRMR, and high CFI.

### Comparing Male and Female Factor Structures

Models of PTS in males and females were compared using local standardized root mean-square difference (localSRMD) (19). Briefly, localSRMD indexes the extent to which each parameter in the model (i.e., factor loadings) differs across groups, here defined as male and female. To compare across sex, a covariance matrix was constructed using all PTS indicators from both sexes (twelve total indicators) and subsequently used to estimate unconstrained (all indicators are estimated freely) and constrained models (the indicators we compare across models were forced to be equal). LocalSRMD values are interpretable similar to Cohen’s *d*, so localSRMD >0.1 and <0.3 indicated a small difference between models, localSRMD >0.3 and <0.5 indicated a medium difference between models, and local SRMD >0.5 indicated a large difference between models. LocalSRMD < 0.1 indicated a trivial difference. A χ^2^ nested model comparison was performed to test the exact equivalence of parameters by sex. A nontrivial localSRMD and significant χ^2^ (p<0.05) must both be true to render the models appreciably different across sex.

### Annotating PTS Factor GWAS

GWAS for PTS factors 1 (PTS-f_1_) and 2 (PTS-f_2_) were annotated using Multi-marker Analysis of GenoMic Annotation (MAGMA v1.08) implemented in FUMA v1.5.3 using 2-kb positional mapping around each lead SNP. Linkage disequilibrium independent loci were defined by lead SNPs with p<5×10^-8^ and surrounding SNPs with *r*^*2*^>0.6 with the lead SNP. Enrichment of tissue transcriptomic profiles was tested using to GTEx v8 for investigation of 54 general tissue types and BrainSpan for investigation of developmental stage and age of brain tissues. Multiple testing correction was applied using FDR (5%) considering all tissue-types or cell-types tested in both sexes. Genome-wide significant SNPs had association p<5×10^-8^ while genome-wide significant genes had association p<2.58×10^-6^ based on a Bonferroni correction for the 19,365 protein-coding genes mapped by FUMA-MAGMA. Significant SNPs and genes for each PTS factor were subjected to phenome-wide association testing in the GWAS Atlas PheWAS browser. Enrichment of associated phenotypes was determined using hypergeometric tests of 4,756 traits separated into 25 trait domains: activities (N=137), aging (N=5), body structures (N=18), cardiovascular (N=162), cellular (N=1,143), cognitive (N=78), connective tissue (N=14), dermatological (N=31), ear, nose, and throat (N=6), endocrine (N=67), environmental (N=92), gastrointestinal (N=42), hematological (N=38), immunological (N=365), infection (N=3), metabolic (N=1,259), mortality (N=46), muscular (N=5), neoplasms (N=49), neurological (N=437), nutritional (N=46), ophthalmological (N=28), psychiatric (N=321), reproduction (N=76), respiratory (N=74), skeletal (N=194), and social interactions (N=20).

### Linkage Disequilibrium Score Regression (LDSC)

LDSC was used to estimate the *h*^*2*^-SNP of each PTS-factor in each sex based on the 1000 Genomes Project European ancestry reference panel. The major histocompatibility complex region was excluded from these analyses due to its complex linkage disequilibrium structure. LDSC also was used to estimate cross-trait genetic correlation (*r*_*g*_) with respect to 523 UKB traits with suitably powerful *h*^*2*^-SNP estimates in both males and females (*h*^*2*^-SNP Z-score>4 in both sexes). Two-sided Z-tests were used to compare *r*_*g*_ estimates between the two factors within each sex. Traits correlated with one factor (FDR<0.05) but not with the other factor (P>0.05) *and* with a significant difference between r_g_ were considered”specific” to that factor.

### Latent Causal Variable Modeling

The Latent Causal Variable (LCV) method was used to identify putative causal relationships between PTS-f_1_ and PTS-f_2_ and each phenotype from the UKB. Briefly, LCV distinguishes genetic correlation from causation by modeling a latent mediator with a causal effect on each trait (20). All genetic causality proportions (gĉps) between two traits were estimated with the PTS factor as trait 1 and the UKB trait as trait 2. With this coding in mind, the posterior mean gĉps reported here are interpreted as follows: (i) |gĉp| between 0.7-1 indicates a fully causal relationship, (ii) gĉp>0 indicates that PTS causes trait 2, (iii) gĉp<0 indicates that PTS is caused by trait 2, and (iv) the causal effect (i.e.,”increase” or”decrease” in risk) for significant gĉps were deduced from the genetic correlation estimate (21,22). Multiple testing correction was performed using a false discovery rate of 5%. Assessing all traits at an LCV two-tailed p<0.05, hypergeometric testing was performed to determine enrichment of putatively causal trait domains based on six relevant assessments of the data: (i) PTS-f_1_ and PTS-f_2_ cause trait 2; (ii) trait 2 causes PTS-f_1_ and PTS-f_2_; (iii) all traits with significant putative causal effects with PTS-f_1_ and PTS-f_2_ but insignificant gĉp differences measured by two-sided Z-tests, herein listed as”concordant”; (iv) traits with significant gĉps relative to one PTS factor and not the other, and significant gĉp differences measured by two-sided Z-tests, herein listed as”discordant”; (v) PTS-f_1_ specific; (vi) PTS-f_2_ specific. Two-sided Z-tests were used to compare gĉp estimates between the two factors within each sex.

## RESULTS

### Structure of PTS factors in males and females

Using GenomicSEM, common factor and two-factor models were explored using posttraumatic stress symptoms in males and females. EFAs suggested two-factor models fit best in both males (cumulative variance explained for 1-factor model = 0.762, 2-factor model = 0.914) and females (cumulative variance explained for 1-factor model = 0.796, 2-factor model = 0.887; Table S2). Models of male and female PTS symptoms differed substantially (localSRMD = 3.12) and significantly (χ^2^(5) = 28.03, p = 3.6×10^-5^). The unstandardized loading for the *thoughts* indicator differed most between males and females. Freeing this parameter reduced localSRMD to 2.30, which still indicated a large difference between sexes, but the difference was no longer significant by χ^2^ testing (χ^2^(4) = 9.44, p = 0.051). With the relaxed model (i.e., freeing the *thoughts* indicator) we established partial invariance of the factors, but were underpowered to detect sex differences in loadings for specific indicators of PTS-f_1_ and f_2_. However, there were noteworthy qualitative patterns that differed by sex. Male PTS-f_1_ largely reflected *irritability* (loading=1, se=0.417, p=0.015) and *feeling distant* (loading=0.95, se=0.233, p=0.001) while PTS-f_2_ represented *upset feelings* (loading=0.938, se=0.462, p=0.042). Female PTS-f1 reflected *trouble concentrating* (loading=0.995, se=0.096, p=3.72×10^-25^), *avoidance* (loading=0.957, se=0.091, p=4.17×10^-26^), and *feeling distant* (loading=0.904, se=0.120, p=4.68×10^-14^) while female PTS-f_2_ represented *irritability* (loading=0.954, se=0.258, p=2.16×10^-4^). CFAs were constructed for all four models (Table S3) and support the separate two-factor constructions for both males and females (Figure 1, Figure S1, and Table S4). These models were used for GWAS of each factor.

**Figure 1.**
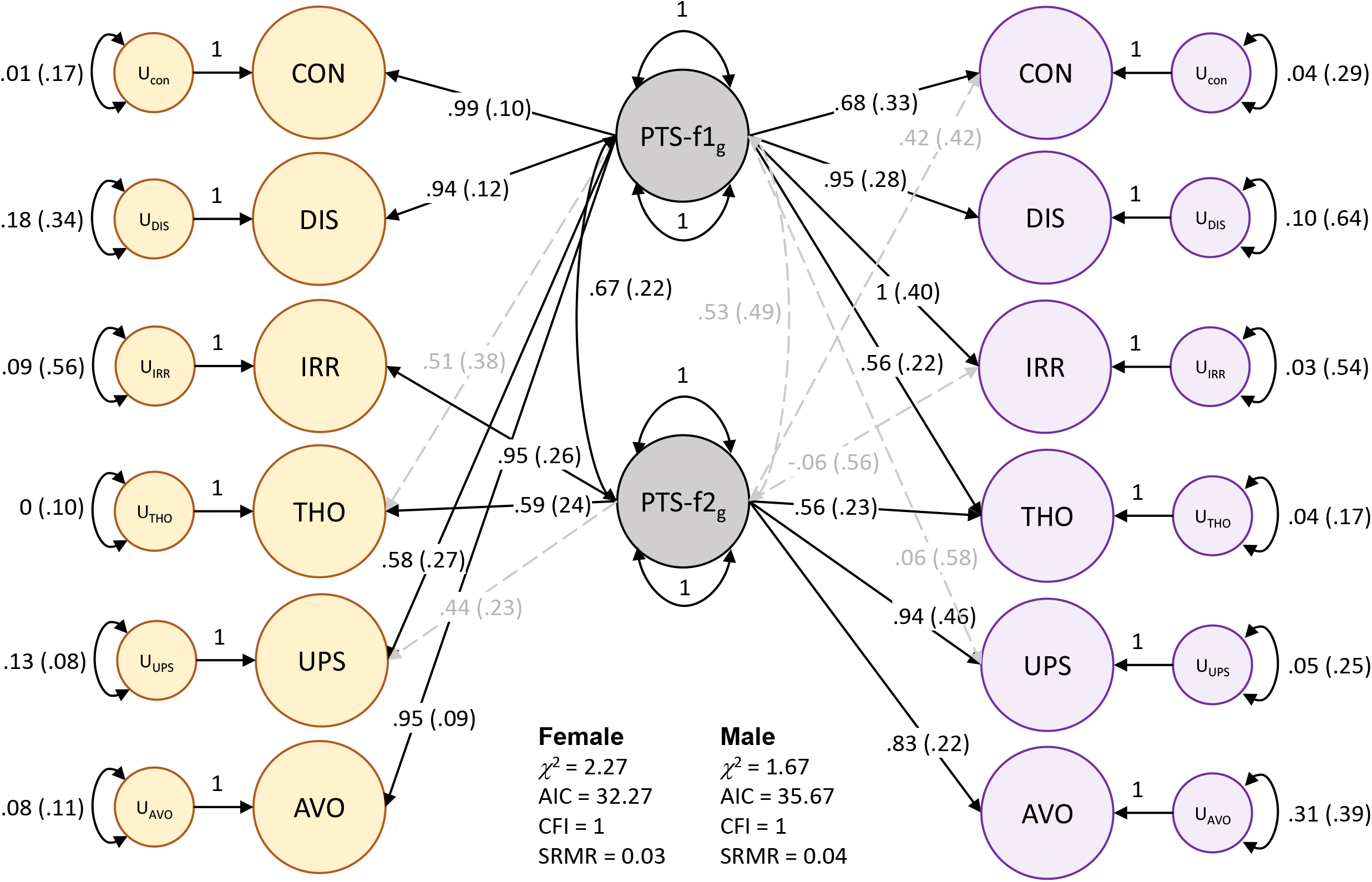
Two-factor model of posttraumatic stress in females (yellow) and males (purple). Standardized loadings are shown for each indicator: CON = trouble concentrating, DIS = feeling distant from others, IRR = irritability, THO = recent distributing thoughts, UPS = recent upset feelings, AVO = avoidance. Significant factor loadings (p<0.05) are shown in solid black lines while dashed grey lines show loadings suggested by EFA but did not reach statistical significance (p>0.05) in CFA.

**Figure 2.**
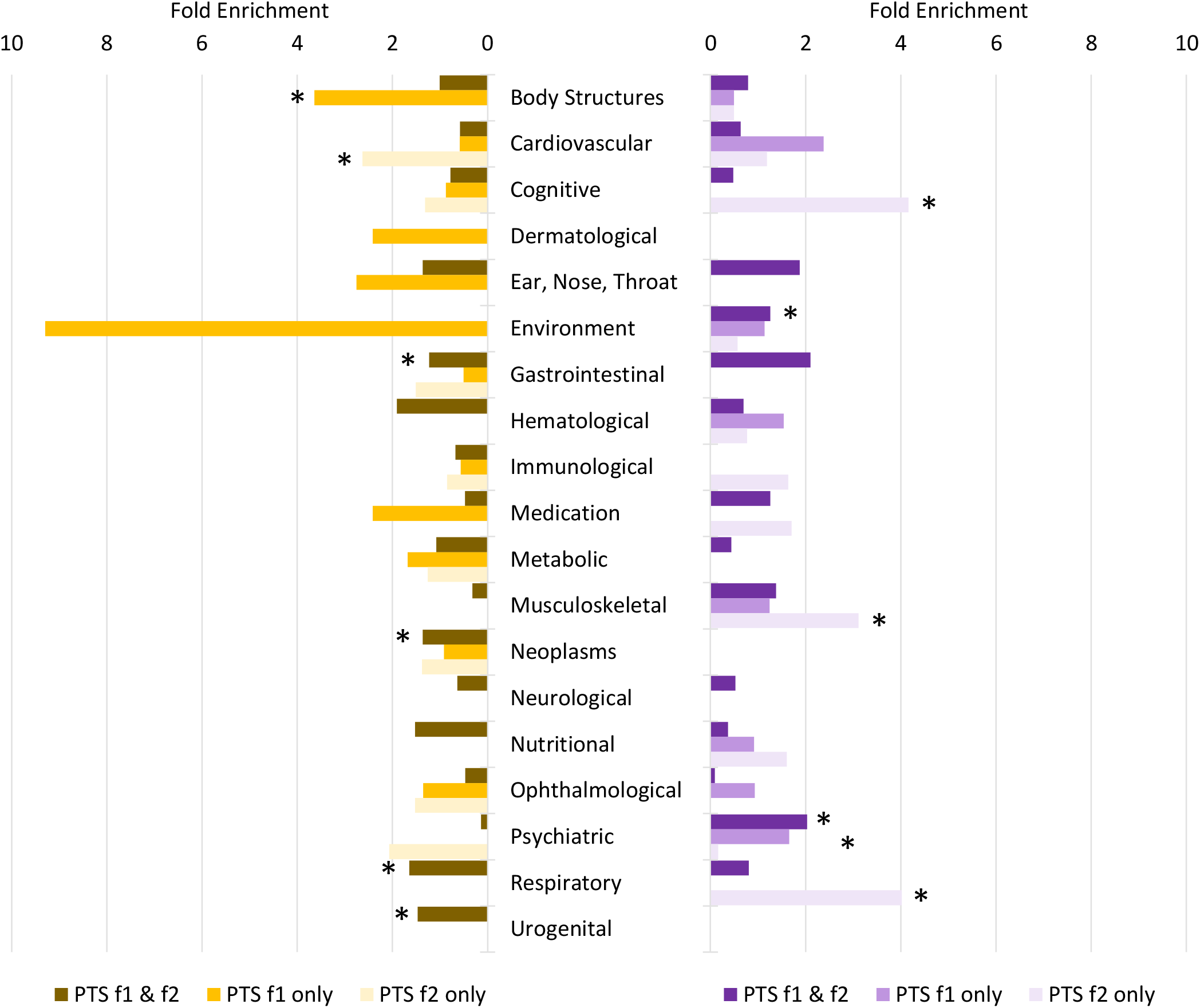
Patterns of trait domain enrichment among genetic correlates of male (purple) and female (yellow) posttraumatic stress (PTS) factors. Fold enrichments were calculated using hypergeometric tests (Table S12) with asterisks indicating significant enrichment (P<0.05).

**Figure 3.**
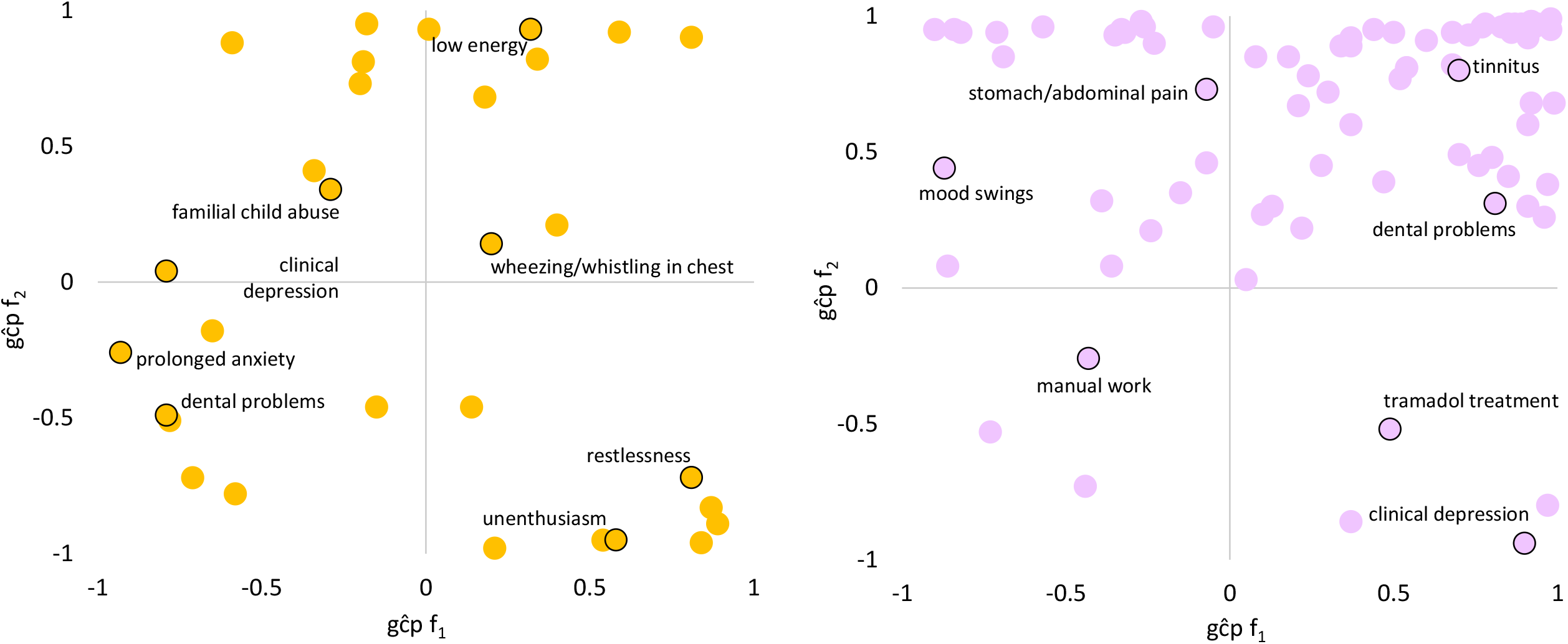
Patterns of significant trait domain genetic causality proportions (gĉps) from latent causal variable analysis of female (left) and male (right). Top-right: caused by f_1_, caused by f_2_. Top-left: causes f_1_, caused by f_2_. Bottom-right: caused by f_1_, causes f_2_. Bottom-left: causes f_1_, causes f_2_. Select traits are highlighted in each quadrant. All gĉp results are shown in Tables S13 and S14.

### Genetic architecture of male and female PTS factors

Each PTS factor from males and females had non-zero SNP-heritability estimates: male PTS-f_1_ *h*^*2*^-SNP=1.85%, se=0.33, male PTS-f_2_ *h*^*2*^-SNP =1.47%, se=0.32, female PTS-f_1_ *h*^*2*^-SNP =3.87%, se=0.43; female PTS-f_2_ *h*^*2*^-SNP =3.53%, se=0.37. There were two loci associated with female PTS-f_1_: (i) rs146918648 (β=-0.072, p=6.73×10^-9^) positionally mapped to an intronic region of *SCAND3*, previously detected in studies of well-being, depressive symptoms, and positive affect (Tables S5 and S6) and (ii) rs72813410-C (β=-0.08, P=1.94×10^-8^) which positionally mapped to a intronic region of *WDPCP*, previously detected in studies of smoking initiation and age of first sexual intercourse (Tables S7 and S8). Gene-based association testing of female PTS-f_1_ revealed an association with *FAM120A* (Z=4.56, P=2.52×10^-6^), which has been detected previously in studies of neuroticism, loneliness, and comparative body size at age 10 (Table S9). The GWAS for female PTS-f_1_ was enriched for transcriptomic effects from spinal cord cervical c-1 (β=0.015, p=0.035), hippocampus (β=0.013, p=0.039), hypothalamus (β=0.013, p=0.040), and substantia nigra (β=0.013, p=0.049; Table S10), while the GWAS of female PTS-f_2_ was enriched for vaginal transcriptomic effects (β=0.018, p=0.046).

There were no genome-wide significant loci associated with either male PTS factor (which showed lower heritabilities than the female factors), but the genome-wide signal of male PTS-f_2_ was weakly enriched for cerebellar (β=0.009, p=0.049), aortic (β=0.018, p=0.024), and tibial artery transcriptomic signatures (β=0.021, p=0.008; Table S10).

### Genetic Correlation

After multiple testing correction (FDR<0.05), 199 of 523 traits were genetically correlated with male PTS-f_1_ and PTS-f_2_, respectively (Table S11). These were enriched for psychiatric traits (2.03-fold, p=1.29×10^-14^; Table S12). There were 10 traits associated with male PTS-f_1_ (FDR<0.05) but not associated with male PTS-f_2_ (P>0.05). The strongest of these was with *number of full sisters* (UKB Field ID 1883; *r*_*g*_ with male PTS-f_1_=0.439, p=0.003 and with male PTS-f_2_=0.238, P=0.133). There were 28 traits associated with male PTS-f_2_ (FDR<0.05) but not associated with male PTS-f_1_ (p>0.05). These were enriched for cognitive (4.66-fold, p=4.34×10^-4^), musculoskeletal (2.66-fold, p=0.022), and nutritional (2.85-fold, p=0.016) trait domains. The strongest of these was with *fluid intelligence* (*positional arithmetic*, UKB Field ID 4968; *r*_*g*_ with male PTS-f_1_=-0.175, p=0.160 and with male PTS-f_2_=-0.477, p=0.002).

After multiple testing correction (FDR<0.05), there were 274 traits genetically correlated with female PTS-f_1_ and PTS-f_2_ (Table S11). These were enriched for respiratory (1.64-fold, p=9.29×10^-12^), gastrointestinal (1.23-fold, p=0.001), and urogenital (1.49-fold, p=0.047) trait domains (Table S12). This set of traits was also depleted for psychiatric traits (7.34-fold, p=2.31×10^-7^). There were 29 traits associated with female PTS-f_1_ (FDR<0.05) but not associated with female PTS-f_2_ (p>0.05). These traits were enriched for body structures (3.64-fold, p=9.67×10^-5^) – the strongest result from this trait domain was *whole body water mass* (UKB Field ID 23102; *r*_*g*_ with female PTS-f_1_=0.128, p=5.13×10^-4^ and with female PTS-f_2_=0.074, p=0.056). There were 18 traits associated with female PTS-f_2_ (FDR<0.05) but not associated with female PTS-f_1_ (p>0.05). These were enriched for cardiovascular traits (3.64-fold, p=0.036) and ophthalmological traits (8.50-fold, p=0.018) – the strongest genetically correlated trait from these domains was *coronary atherosclerosis* (UKB Field ID I9_CORATHER; *r*_*g*_ with female PTS-f_1_=0.146, p=0.126 and with female PTS-f_2_=0.219, P=0.023).

### Putative Causal Relationships

LCV was used to determine if genetic correlations suggested a putatively causal effect mediated by a latent variable. After multiple testing correction, there were many putative causal relationships with each PTS factor (male PTS-f_1_ = 99; male PTS-f_2_ = 166; female PTS-f_1_ = 34; female PTS-f_2_ = 80). In males and females, nearly all analysis strata were enriched for putative causal relationships with psychiatric traits (Tables S13-S15); however, male PTS-f_2_-specific results were depleted for psychiatric traits (3.42-fold depletion, p=8.83×10^-7^). Male PTS-f_1_-specific traits were enriched for traits related to medication intake (3.73-fold, p=0.032) while male PTS-f_2_-specific traits were enriched for body structure (2.84-fold, p=5.51×10^-7^) and cognitive (3.22-fold, p=0.009) causal relationships. In females, PTS-f_1_-specific traits were enriched in the metabolic domain (28.08-fold enrichment, p=0.035).

Of the medication-related traits specifically enriched in male PTS-f_1_, we observed conflicting putative causal estimates with respect to omeprazole use. General omeprazole use was putatively causal for male PTS-f_1_ (gĉp=-0.05, p=0.005); however, omeprazole use ascertained in the context of pain relief, constipation, and/or heartburn was caused by PTS-f_1_ at much larger magnitude (gĉp=0.39, p=0.020). The body structure traits with putative causal effects relative to male PTS-f_2_ were mainly for measures of body fat percentages (including whole body, arms, legs, and waist), as well as body mass index (BMI; gĉp=0.8; p=3.41×10^-8^) increasing as a result of PTS-f_2_. Additionally, in the cognitive domain, PTS-f_2_ had causal effects with decreased performance on fluid intelligence (gĉp=0.56; p=1.53×10^-6^) and reaction time (gĉp=0.89; p=6.24×10^-10^) tests. The most significant male PTS-f_2_-specific findings indicated that PTS-f_2_ causes repeated disturbing thoughts of stressful experiences (gĉp=0.9; p=6.24×10^-10^) and increased risk-taking behaviour (gĉp=0.44; p=2.48×10^-8^). In females, enrichment was detected for metabolic traits in PTS-f_1_, in which the PTS-f_1_-specific association was high cholesterol having a causal effect on PTS-f_1_ (gĉp=-0.78; p=1.04×10^-7^). Female PTS-f_2_ had many specific putative causal relations with psychiatric traits; the most significant of which were anxiety and depression-related: PTS-f_2_ causing nervous feelings (gĉp=-0.53; p=2.16×10^-32^) and extended periods of unenthusiasm/disinterest (gĉp=-0.95; p=3.13×10^-30^) and tiredness/lethargy causing PTS-f_2_ (gĉp=-0.91; p=5.09×10^-21^).

## DISCUSSION

Through multivariate modelling of PTSD symptom GWAS and assessment of genetic correlates and putative causal phenotypes related to each symptom factor, our study identified patterns of phenotypic differences which could inform sex differences in PTSD etiology. Consistent with prior literature, female PTS factors had slightly higher SNP-based *h*^*2*^ estimates than male PTS factors. Looking at factor structure, male PTS factors were uncorrelated while females PTS factors were correlated. These findings may in part be attributed to women reporting more severe PTS symptoms relative to men. Our novel approach to compare male and female factor structures in UKB indicated that differences in factor loadings of PTSD symptoms do exist across males and females. Though model comparisons support these differences, we were underpowered to identify which factors differ (16). However, the pattern of indicators that load onto each factor show genetic differences between sexes that are reinforced among phenotype-level factor modeling. Overstreet et al. demonstrate among military veterans (92.2% male) that irritability and difficulty concentrating cluster together in a dysphoric arousal factor (16). Despite using a civilian cohort to measure PTSD symptoms, our genetic investigation supports this clustering. However, we showed using genetic data that irritability and difficulty concentrating significantly and strongly load onto distinct factors. Below we detail findings from GWAS of each PTS factor to understand what each latent feature of PTS represents in the context of health and disease relevance.

Across all four GWAS (two each in males and females), we identified two genome-wide significant loci associated with female f_1_: *SCAND3* (SCAN domain containing 3) and *WDPCP* (WD repeat containing planar cell polarity effector). SCAND3 was previously identified as potentially diagnostic for hepatocellular carcinoma, a cancer of the liver (23). As cancer may serve as an index trauma, SCAND3 may serve as an indicator of PTS severity, though this relationship requires dedicated testing (24–26). We identified *WDPCP* (rs72813410-C) as a risk decreasing variant for female PTS-f_1_. This gene was previously identified as a risk-increasing genome-wide significant finding in a cross-disorder study of the impulsivity-compulsivity spectrum (comprised of Tourette Syndrome, obsessive compulsive disorder, ADHD, and autism spectrum disorder) (27). WDPCP also associated with DNA methylation in past child soldiers (28) and tinnitus prevalence (29). These findings, along with lack of enrichment for psychiatric genetic correlates and enrichment of putatively causal relationships with body structures, support the identity of female PTS-f_1_ as a general factor of non-psychiatric disease/disorder risk. Despite lack of individual significant loci identified in the GWAS of female PTS-f_2_, male PTS-f_1_, and male PTS-f_2_, we detail their genetically correlated and putatively causal profiles below.

Female PTS-f_2_ was enriched for cardiovascular genetic correlates and reinforced well-studied relationships between cardiovascular health and PTSD (30). Though correlated, these effects were not detected by enrichment tests of putatively causal trait relationships. We attribute this lack of enrichment to the also well documented difficulties in untangling the relationship between PTSD and cardiovascular disease. Briefly, despite evidence of causal effects measured by Mendelian randomization (30), bidirectional effects persisted with certain cardiovascular phenotypes. In one attempt to resolve these effects, the onset of PTSD relative to onset of cardiovascular disease appeared to play some role in the observed bidirectional relationship between these traits (31).

Male PTS-f_1_ and PTS-f_2_ were genetically correlated with psychiatric traits. Male PTS-f_1_ was not enriched for any genetic correlates but did have specific enrichment of putative causal effects with medication use traits localized to pain relief medications: tramadol (a synthetic opioid), omeprazole (a proton pump inhibitor), and aspirin. Low-dose aspirin is often used for the prevention of myocardial infarction, as well as an over-the-counter analgesic commonly taken by PTSD patients (32–34). Patients with PTSD report higher use of opioid and non-opioid analgesics relative to non-PTSD patients (35). The relationship between opioid use and PTSD is complex and multidirectional, as opioids may be used to address pain caused by PTSD, while also potentially being related to the traumatic event(s) leading to PTSD. Our putative causal estimates between pain medication and PTS using the LCV approach were bidirectional and consistent with literature reports of chronic pain and internalizing disorders such as depression (36). These findings reinforce several scenarios of their possible causal relationship including: an index trauma causes chronic pain and leads to elevated PTS, (ii) chronic pain (e.g., lower back pain, migraines) itself is a traumatic experience contributing to PTS, and (iii) intrusive and hyperarousal symptoms cause pain complaints and/or pain catastrophizing, respectively (37). These relationships are largely considered evidence of shared vulnerability and are reviewed in greater detail by Asmundson et al. (38). Male PTS-f_2_ was enriched for musculoskeletal traits at the level of genetic correlation and putative causality. Unlike male PTS-f_1_, these results highlight effects of stress on dental health and mid-to-low back pain (39,40). In sum, male PTS findings support the role of pain in PTS. There appears to be pain area-specific and factor-specific effects, but these require further investigation to verify.

LCV analyses uncovered several sex-based differences in PTSD biology, etiology, and symptomatology. While it was expected for psychiatric enrichment to be present in both sexes, the observations within this trait domain differ. In males, for example, the most significant concordant traits were tiredness/lethargy, trouble relaxing, and low energy, potentially corresponding to depression, anxiety, and sleep-related symptoms. In females, the most significant concordant traits were suffering from nerves, familial abuse as a child, and decreased ability to confide. These findings in females correspond not only to anxiety-related traits, but also treatment, coping mechanisms, and trauma exposure. The genetic causal effects related to familial abuse provide support for studies showing that greater genetic liability for PTSD in women may contribute to earlier onset and/or more severe symptoms, as well as women more generally experiencing trauma early in life (41). Further, we found that many of the significantly associated traits in women were anxiety-related (e.g. suffering from nerves, extended periods of anxiety, tenseness/restlessness were associated with both PTS-f_1_ and PTS-f_2_), supporting findings that women are more prone to anxious symptoms of PTSD (42,43). These findings may also simply reflect the increased prevalence of anxiety disorders in women, as well as the strong comorbidity between PTSD and anxiety disorders (44,45). Further study of female- and male-specific disorder and symptom factor structures may aid in untangling these observations.

Our study has three primary limitations to consider. First, the UKB dataset primarily represents European ancestry individuals of a generally higher socioeconomic bracket (46). This means that PTS symptoms are more likely (i) relatively mild due to lower prevalence of exposure to traumatic events and (ii) reflective of civilian trauma patterns; military cohorts have been observed to experience greater resilience to trauma, thereby altering PTSD etiology (47). Our results may therefore only largely apply to a European population and/or to higher socioeconomic status groups. Second, the UKB has information for a diverse collection of adult and childhood events that could be considered traumatic, but there is no information on the quantity or severity of the events. This information may demonstrate that the PTS symptoms reported by UKB participants are enriched for certain types of experiences. PTS patterns following specific traumas or trauma domains therefore warrant dedicated investigation; however, biobank-type datasets are required to achieve adequate power for these kinds of studies, and this level of phenotype detail is rare in biobank contexts. Finally, on a statistical level, indicator GWAS used to estimate PTS factor structure fit a linear model to ordinally distributed data. With sample sizes as large as the UKB, this mismatch of model suitability may not be able to control type I error rate for rarer variants (48). Therefore, the genetic signal of the PTS indicators used here may not fully reflect their true polygenic nature.

In summary, we demonstrate different PTS genetic factor structures between sexes, highlighting the importance of explicitly considering sex in genetic studies of PTSD. By modeling genetic correlation and putative causal relationships of each factor in males and females, we present preliminary data to inform how different dimensions of PTS may influence other traits, behaviors, and disorders. These data inform the etiological differences between males and females supported by existing literature and introduce new hypotheses to help understand the biology of PTS and PTSD.

## ACKNOWLEDGEMENTS

This project is partially supported by funding from the University of Toronto Data Sciences Institute.

## DISCLOSURES

Dr. Stein has in the past 3 years received consulting income from Acadia Pharmaceuticals, Aptinyx, atai Life Sciences, BigHealth, Biogen, Bionomics, BioXcel Therapeutics, Boehringer Ingelheim, Clexio, Eisai, EmpowerPharm, Engrail Therapeutics, Janssen, Jazz Pharmaceuticals, NeuroTrauma Sciences, PureTech Health, Sage Therapeutics, Sumitomo Pharma, and Roche/Genentech. Dr. Stein has stock options in Oxeia Biopharmaceuticals and EpiVario. He has been paid for his editorial work on *Depression and Anxiety* (Editor-in-Chief), *Biological Psychiatry* (Deputy Editor), and *UpToDate* (Co-Editor-in-Chief for Psychiatry).

Dr. Gelernter is paid for editorial work (*Complex Psychiatry*).

The other authors have no competing interests to report.

## AUTHOR CONTRIBUTIONS

FRW and AM designed the project. FRW and AM collected the data and performed all analyses. AM, MBS, JG, and FRW interpreted the analyses. AM and FRW wrote the paper. AM, MBS, JG, and FRW edited the paper. FRW supervised the study.

## DATA AVAILABILITY

PTS factor summary statistics can be accessed via Zenodo: 10.5281/zenodo.8061505.

## Notes

### Author Declarations

The study used GWAS summary data available for download from http://www.nealelab.is/uk-biobank.

