## Supplementary material for "Sex-stratified genomic structural equation models of posttraumatic stress inform PTSD etiology": Supp_Material

### **SUPPLEMENTARY FIGURES**

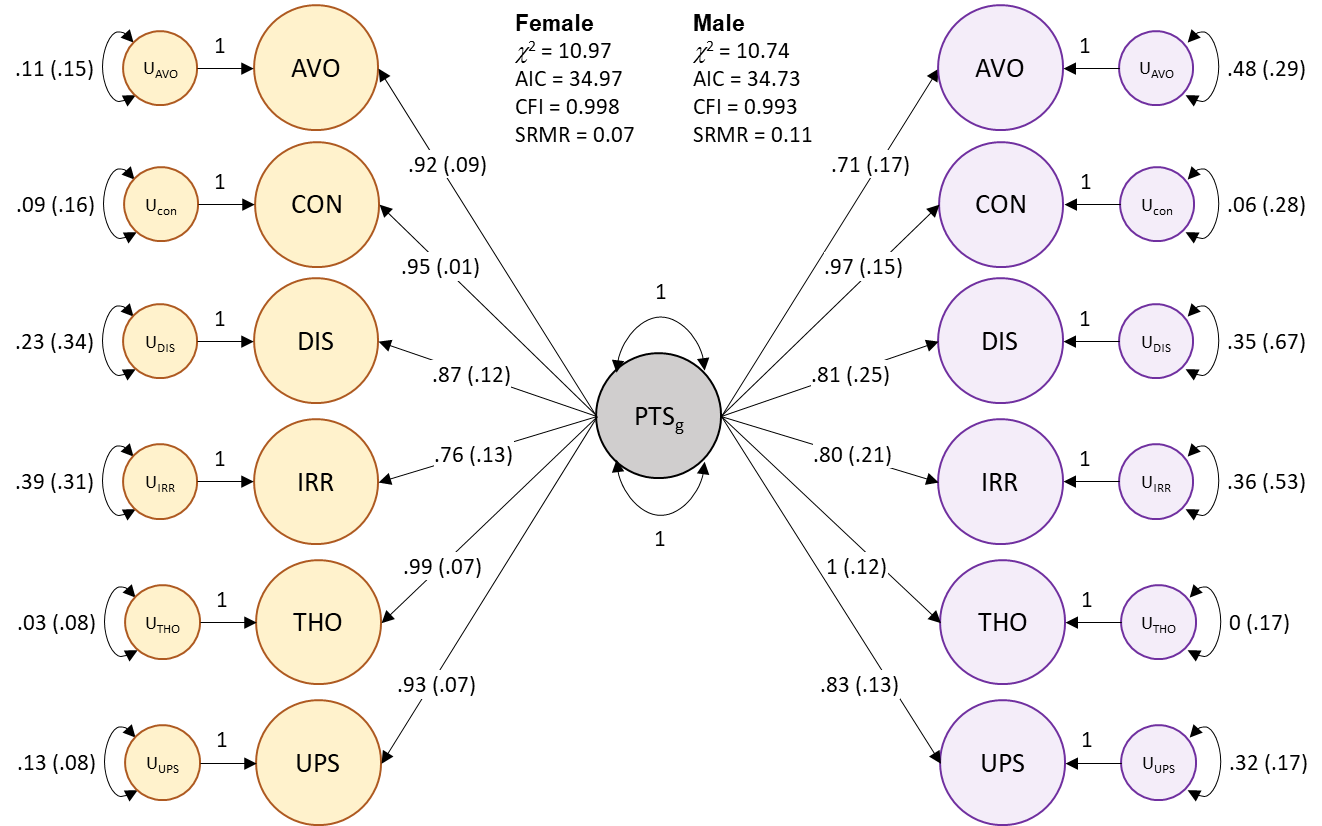

**Figure S1.** Common factor model of posttraumatic stress in females (yellow) and males (purple). Standardized loadings are shown for each indicator: CON = trouble concentrating, DIS = feeling distant from others, IRR = irritability, THO = recent distributing thoughts, UPS = recent upset feelings, AVO = avoidance. All factor loadings were significant (p<0.05).

### **SUPPLEMENTARY TABLES**

**Table S1.** Trait description and observed-scale heritability estimates for each posttraumatic stress indicator. Heritability comparisons between males and females were calculated using two-sided Z-tests.

**Table S2.** One- and two-factor exploratory factor analysis results for posttraumatic stress indicators. Highlighted traits were included in confirmatory factor analyses.

**Table S3.** Confirmatory factor fit statistics for each factor structure tested. CFI = comparative fit statistics, AIC = Akaike information criterion, SRMR = standardized root mean square residual.

**Table S4.** Loading values for each indicator on the two-factor model stratified by sex.

**Table S5.** GWAS Atlas PheWAS results for female PTS-f_1_ locus rs146918648.

**Table S6.** GWAS Atlas PheWAS results for female PTS-f_1_ locus *SCAND3*.

**Table S7.** GWAS Atlas PheWAS results for female PTS-f_1_ locus rs72813410.

**Table S8.** GWAS Atlas PheWAS results for female PTS-f_1_ locus *WDPCP*.

**Table S9.** GWAS Atlas PheWAS results for female PTS-f_1_ locus *FAM120A*.

**Table S10.** Enrichment of brain transcriptomic profiles in the GWAS for female PTS-f_1_. Highlighted tissues are nominally significant (p<0.05).

**Table S11.** Genetic correlation estimates between each PTS factor and a UK Biobank trait with a heritability Z-score > 4. Comparisons between factors (within each sex) were performed using two-sided Z-tests.

**Table S12.** Results of hypergeometric enrichment tests for genetic correlation results (Table S11) for traits associated with both factors and traits uniquely associated with one trait. Highlighted trait domains are significantly enriched.

**Table S13.** Genetic causality proportion estimates between male PTS factors and a UK Biobank trait. Comparisons between factors (within each sex) were performed using two-sided Z-tests.

**Table S14.** Genetic causality proportion estimates between female PTS factors and a UK Biobank trait. Comparisons between factors (within each sex) were performed using two-sided Z-tests.

**Table S15.** Results of hypergeometric enrichment tests for LCV results for all categories (PTS-f_1_ and PTS-f_2_ cause trait 2, trait 2 causes PTS-f_1_ and PTS-f_2_, concordant, discordant, PTS-f_1_-specific, PTS-f_2_-specific). Highlighted trait domains are significantly enriched.
